# Unmet Therapeutic Needs as Drivers of Drug Therapy Problems among Ethiopian Cardiovascular Outpatients: A Cross-Sectional Study

**DOI:** 10.1101/2025.08.04.25332971

**Authors:** Hailu Chare Koyra, Eshetu Ayele Alaro

## Abstract

**Background:** Drug therapy problems (DTPs) defined as medication-related events compromising optimal health outcomes, present particular challenges for patients with cardiovascular disease (CVD) due to complex regimens for multiple comorbidities, often resulting in adverse clinical consequences. This hospital-based cross-sectional study assessed the prevalence and determinants of DTPs among adult ambulatory CVD patients at Wolaita Sodo University Comprehensive Specialized Hospital from May to July 2022.

**Methods:** We conducted an institution-based cross-sectional study of ambulatory cardiovascular disease (CVD) patients receiving chronic care at Wolaita Sodo University Comprehensive Specialized Hospital. Data were collected through structured patient interviews and comprehensive medical record reviews. Drug therapy problems (DTPs) were systematically identified and classified using Cipolle’s validated methodology, adapted for the local patient population. We employed binary logistic regression to assess individual associations between predictor variables and DTPs, followed by multiple logistic regression to control for potential confounders. Statistical significance was determined at p≤0.05, with 95% confidence intervals used to estimate precision of effect measures.

**Result:** Among 195 ambulatory CVD patients, hypertension (59.5%, n=116) and heart failure (47.2%, n=92) were the most prevalent conditions. The study identified drug therapy problems (DTPs) in 69.7% of patients (n=136), totalling 168 DTPs (mean 1.3 SD 0.461 per affected patient). Unmet therapeutic needs (41.0%) and medication non-adherence (24.4%) emerged as the most common DTP categories. ASCVD risk assessment classified 40% of evaluated patients as high-risk (10-year risk ≥20%). Significant predictors of DTPs included polypharmacy (AOR=3.48, 95%CI:1.47-8.24) and age (25-64 years: AOR=16.38), while comorbidities (AOR=0.15) and prior hospitalization (AOR=0.31) demonstrated protective effects.

**Conclusion and recommendation:** This study identifies unmet therapeutic needs and medication non-adherence as the predominant drug therapy problems among ambulatory CVD patients at Wolaita Sodo University Comprehensive Specialized Hospital, calling for systematic medication reviews, pharmacist-led interventions for high-risk groups, and enhanced adherence support programs to optimize treatment outcomes.

## 1. Background

The global rise in cardiovascular illnesses is the outcome of an epidemiologic change that is occurring in every corner of the world, among all races, ethnic groups, and cultures. Cardiovascular diseases are a group of disorders of the heart and blood vessels(1). Cardiovascular diseases are classified as disorder of rhythm, disorder of heart, disorder of coronary and peripheral vascular disease (2).Most of patients with cardiovascular disease can be treated and managed by proper follow and check-up in an outpatient department, though these CVDs accounts for 7-10% of all adult medical admissions in African hospitals, with heart failure accounting for around 3%–7% of them (3).

Pharmacotherapy plays a vital role in lowering cardiovascular disease morbidity and mortality. These benefits, however, are restricted by drug therapy problems, which can reduce a patients’ quality of life, lengthen hospital stays, and increase the overall cost of healthcare (4)(5)(6). Drug therapy problems (DTPs) are defined as any unintended effects encountered by a patient during treatment (6). A drug-therapy problem (DTPs) is defined as an event or circumstance involving drug therapy that actually or potentially interferes with desired health outcomes(8).

A potential problem is not manifest, but if left unresolved, it may lead to drug-related harm to the patient. Drug therapy problems are associated with negative effects on patient outcomes and have the potential to increase the cost of care. According to Robert J. Cipolle text book of pharmaceutical care practice of 3rd edition, DTPs are classified into seven classes, including: Need additional drug therapy, unnecessary drug therapy, ineffective drug, too low or too high dosage, adverse drug reactions, and noncompliance(8). When drug therapy problems are identified, they are resolved by changing products, doses, or by educating the patient on how to maximize the effectiveness of the medication and then a care plan is developed for each patient, including individualized goals of therapy for each medical condition (9).

Worldwide, CVDs continue to remain the primary cause of death. Much money has been spent to combat this illness, both in terms of prevention and therapy. Due to the high number of co-morbidities present in patients with CVDs, it is challenging to concentrate solely on the management of the CVDs because much co-morbidity has an impact on it directly or indirectly. This can lead to DTPs, which calls for comprehensive medication management services that can identify and address drug therapy issues and enhance patient outcomes(9)(10).

DTPs are problem for health care providers, according to studies, and have a significant impact on patient clinical outcomes, leading in higher morbidity and mortality, long hospital stays, and increased health care cost burden on the patient or the government (10)(11).Furthermore, the complex usage of medications in both institutionalized and ambulatory care settings creates an environment favourable to DTPs. Patients with CVDs are also more likely to take several medications, which increases the risk of DTP(10). DTPs are a major source of worry in health care due to rising costs, morbidity, and mortality. Drug-related morbidity and mortality accounted for $177.4 billion of total costs in 2000, with long-term-care admissions accounting for 18% ($32.8 billion) (11) (12).

Currently studies concerning DTPs in patients with adult ambulatory cardiovascular diseases are limited in Ethiopia. Because most of the studies done in Ethiopia concerning DTPs were specific to one of the CVDs. This study is therefore, going to bridge the gap and help identify DTPs with their respective causes in ambulatory CVD patients at chronic care unit. Furthermore, there are no studies conducted in the area. Therefore, the study findings will help in devising strategies to optimize proper drug use among patients with cardiovascular diseases.

The study findings, therefore, will assist in addressing the modifiable risk factors that healthcare providers will address to reduce morbidity and mortality related to DTPs amongst CVD patients. The findings of this study will, in general, have a positive impact on clinical practice among CVD patients. Therefore, this study was aimed to assess the magnitude of DTPs and their contributing factors in the management of patients with CVD in ambulatory care clinic of Wolaita sodo University comprehensive Specialized Hospital (WSUCSH), Southern Ethiopia.

## 2. Methods and Materials

### 2.1 Study design and study setting

A hospital based cross sectional study design was conducted at the ambulatory care clinic in WSUCSH, Wolaita, Ethiopia. WSUCSH is a teaching and referral hospital. It is the largest public hospital in southern Ethiopia which is estimated to have about 3.5–5 million catchment population. Chronic care was one of the specialty units of the hospital which provides medical services for registered, newly diagnosed and referral CVD patients. The clinic contains three nurses and two residents. The clinic had scheduled follow up (from Monday-Friday) for all ambulatory CVD patients. Based on the data from the health information centre of the hospital, about 412 patients with CVD had follow up in chronic care clinic.

### 2.2 Study population and data collection procedure

Consented CVD patients aged ≥ 18 years who had complete medical records and regular follow-up between May 9 to July 8, 2022 were included in the study. Patients were recruited into the study during their appointment for medication refilling. Patients were excluded if they were newly diagnosed with CVD (*<*6 months), seriously ill to complete the interview, unwilling to give consent, and their medical record not complete or available for further review. A sample of 195 was calculated using a single population proportion formula.

Using the following formula: *n= z^2^ap (1-p)/d^2^,*

Where, n – required sample size

z – Confidence level at 95% (standard value =1.960) at □ / 2

p – Prevalence =72% (0.72)

d – Margin of error at 5% (0.05)

Hence n= (1.96)^2^ (0.72) (0.28) / (0.05)^2^ = 310, n=310

According to data obtained from the hospital office of health management information system, the source population were 412; it is less than 10,000. So that correctional formula was used to get the final sample size (nf). Nf= n/1+n/N = n × N / n + N = 310 × 412 / 310 + 412 =177 Where nf= final sample size, N=total source population

The calculated sample size was n = 177 patients. So that, taking 10% of total sample size as non-response rate in to consideration the minimum sample size required for the study was 195. Based on a study conducted at Gebretsadik Shawo General Hospital, Bonga, South west Ethiopia regarding the prevalence of DTP in 2017 GC, the prevalence rate of DTP among CVD patients were found to be 72%. So 72% taken as a previous prevalence rate, to get possible large sample size, confidence level of 95 % was chosen, Margin of error (d) = 5% and 10% of contingency for non-response rate.(13) We approached a total of 195 participants. Patients were interviewed consecutively according to their appointment schedule using the interview questionnaire and their respective medical chart was retrieved using the retrieval checklist. Interview questions included socio-demographics, medication adherence, medication experience and previous medication history assessment. Respective patient’s medical, medication and laboratory records were reviewed. Two clinical pharmacists and two nurses were recruited for data collection and one day training was provided to familiarize them how to retrieve pertinent data from medical record and how to conduct patient interview.

### 2.3. Drug therapy problems identification and assessment

The DTP evaluation tool was prepared based on Cipolle DTP categories with slight modification(14). It is widely accepted in patient cantered text book, which is standardized guideline for pharmacists while practicing pharmaceutical care service and authorized by Ethiopian Hospital Reform implementation Guidelines (EHRIG) and Pharmaceuticals Fund and Supply Agency (PFSA) to be implemented the provision of pharmaceutical care service in Ethiopia hospitals(15)(16). Adverse drug reaction (ADR) was assessed according to Naranjo algorithm – ADR Probability Scale, which is standardized as well as validated instrument that was used to assess patients’ ADR status with a 10-item self-report measure of ADR that contains 10 questions. If total score is ≥ 9 is definite, if 5 to 8 is probable, if 1 to 4 is possible, if ≤ 0 is doubtful respectively. The identification of DTP was based on a review of patients’ medical and medication records, assessment of laboratory investigations, patients’ interview about medication experience, and physical observation *(Figure 1).* Specific information about medication therapies, such as the recommended drug of choice, recommended dosages, frequency of administration, duration of therapy, drug-interactions and adverse drug events were compared based on details from the standard pharmacotherapy textbooks and most updated guidelines like 2021 Ethiopian standard treatment guideline (STG), 2018 American heart association, 2018 European Society of Cardiology (ESC).

**Figure 1:**
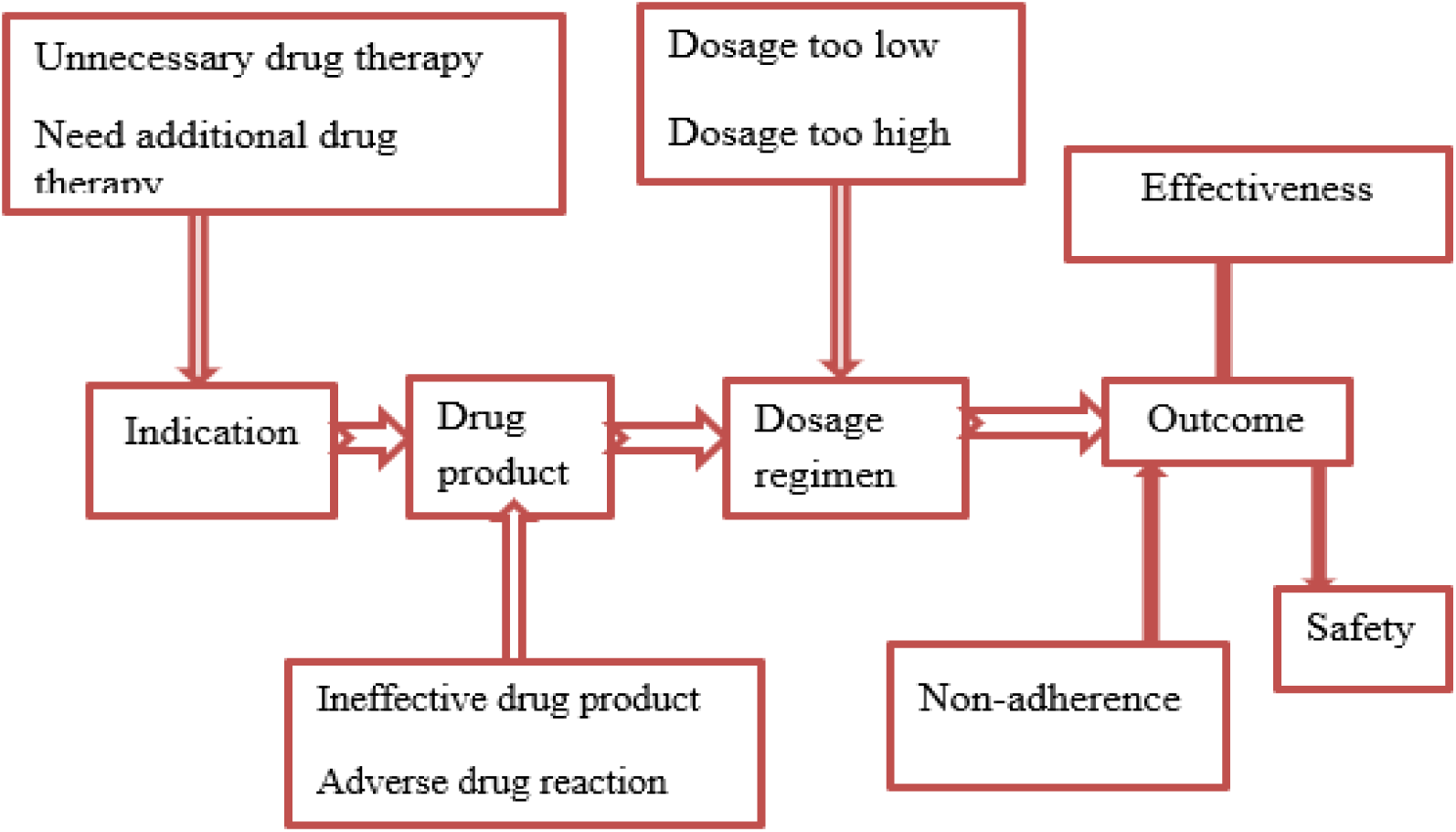
Drug therapy problem assessment and identification sequence.

### 2.4 Definition of terms and variables

**DTP;** refers to any unwanted incident related to medication therapy that actually or potentially affects the desired goals of treatment, which was identified using Cipolle’s method followed by a consensus meeting with experts. DTPs were categorized in to seven major groups’ base on Cipolle’s method. These include: including unnecessary drug therapy, needs additional drug therapy, ineffective drug therapy, dosage too low, adverse drug reaction, dosage too high and noncompliance.

*Category of DTPs were defined and identified based on Cipolle’s method as follows:*

Unnecessary drug therapy is considered when:

- There is no valid medical indication for the drug therapy at this time,
- Multiple drug products are being used for a condition that requires single drug therapy,
- The medical condition is more appropriately treated with nondrug therapy
- The drug therapy is used to treat an avoidable adverse reaction associated with another medication.

Need for additional drug therapy was considered when:

- The medical condition requires the initiation of drug therapy
- Preventive drug therapy is required to reduce the risk of developing a new condition.
- The medical condition requires additional drug therapy to achieve synergistic or additive effects.

Ineffective drug therapy was considered when:

- The least effective drug is used while the most effective drug is available
- The drug is used for medical condition which is refractory to the drug product.
- The drug product used is not an effective product for the medical condition being treated

Dosage too high was considered when:

- The dose is too high
- dosing frequency is too short
- The duration of drug therapy is too long
- The drug interaction occurs that could result in a toxic reaction to the drug product
- The dose of the drug was administered too rapidly

Dosage too low was considered when:

- The dose of the drug is too low to produce the desired response
- The drug interaction occurs that could reduce the amount of active drug available
- The duration of drug therapy is too short to produce the desired response
- The dosage interval is too infrequent to produce the desired response

Adverse drug reaction was considered when:

- The drug causes an undesirable reaction that was not dose-related
- The drug interaction causes an undesirable reaction that is not dose-related
- The drug causes an allergic reaction
- The drug is contraindicated due to risk factors
- The dosage regimen administered or changed too rapidly

Noncompliance is considered if a patient fails to take medications appropriately due to one of the following reasons:

- Lack of understanding the instructions
- Preference not to take the medication
- Forgetfulness
- Inability to swallow or self-administer the drug product appropriately
- Affordability and availability problem

### 2.5. Data analysis

Data were entered into an EPI data management and analysed using SPSS version 20.0. Descriptive analysis was computed as frequency, mean and standard deviation (SD). Multicollinearity was checked to test correlation among predictor variables using variance inflation factor and none was collinear. Univariable logistic regression analysis was performed to determine the association of each independent variable with DTP, and then independent variables with p value <0.25 in univariable analysis were included into a multivariable binary logistic regression model to identify predictors of DTP. A p value of <0.05 was considered statistically significant in all analyses.

### 2.6. Ethical clearance

This study was approved by the institutional review board (IRB) of Wolaita Sodo University, College of Health Sciences with the project No of CHSM/ERC/06/14. The purpose and protocol of this study was fully explained to all study participants. Written informed consent was obtained from all participants included in this study. The privacy of personal information was entirely confidential and protected. All the methods were performed in accordance with approved institutional guidelines.

## 3. Results

### 3.1 Socio-demographic characteristics

In this study, 195 CVD patients were included with the mean age of 54 ± 27.7 ranging from 20 to 110 years. Majority of the patients were in the age group of ≥ 65years (40.5%). More than half of the patients were females (54.4%), while 79.5% and 42.1% of the patients were married and illiterate, respectively. Regarding social drug use behaviour, majority of the patients were coffee users (81.5%). Majority of the patients were followers of and live in rural areas and have no regular monthly income each constituting 69.8%, 69.2% & 35.4 respectively of the study participants. The highest percentage of patients was farmer (42.6%) and around 160(82.1%) patients *(Table 1)*.

**Table 1:**
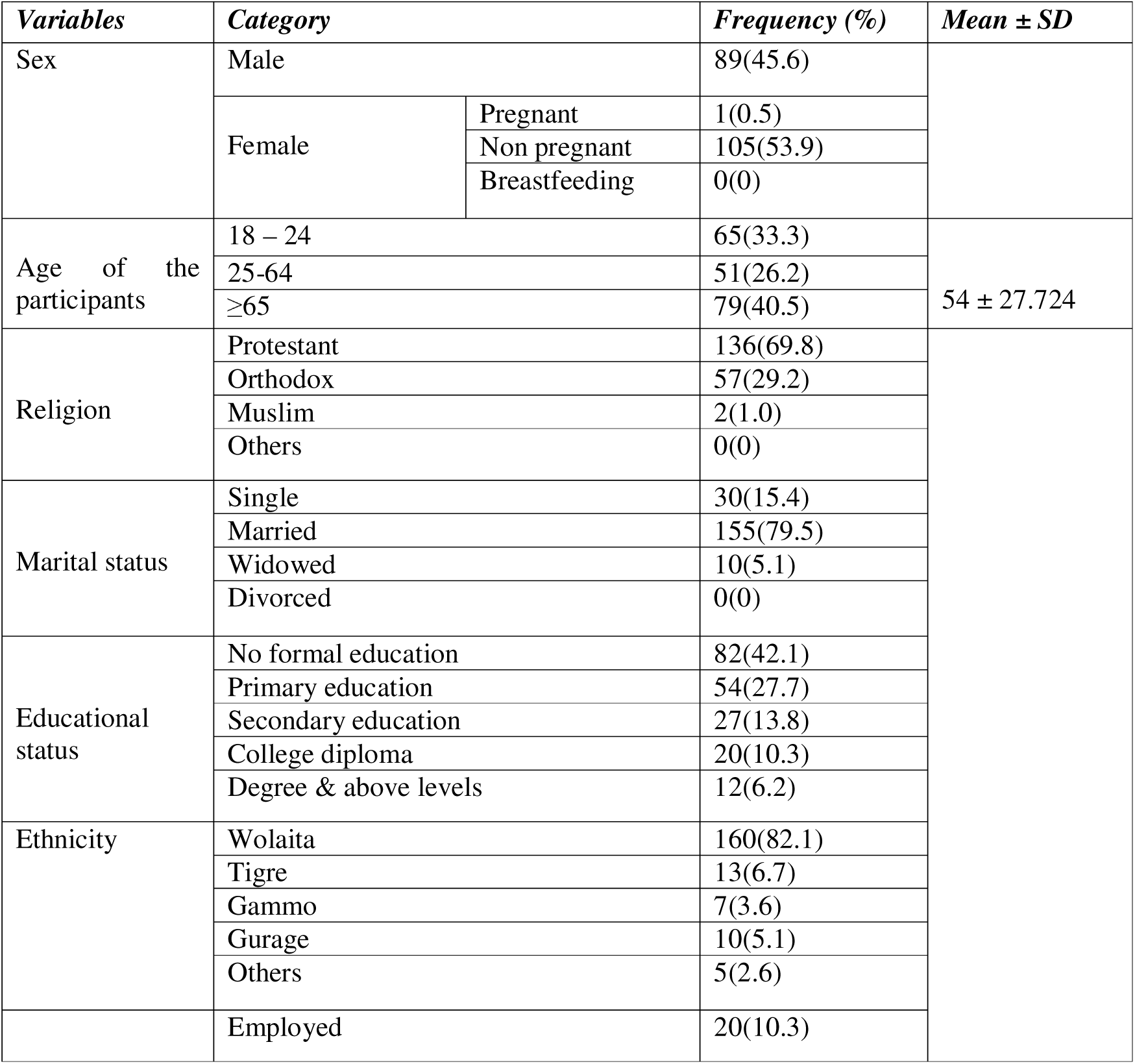
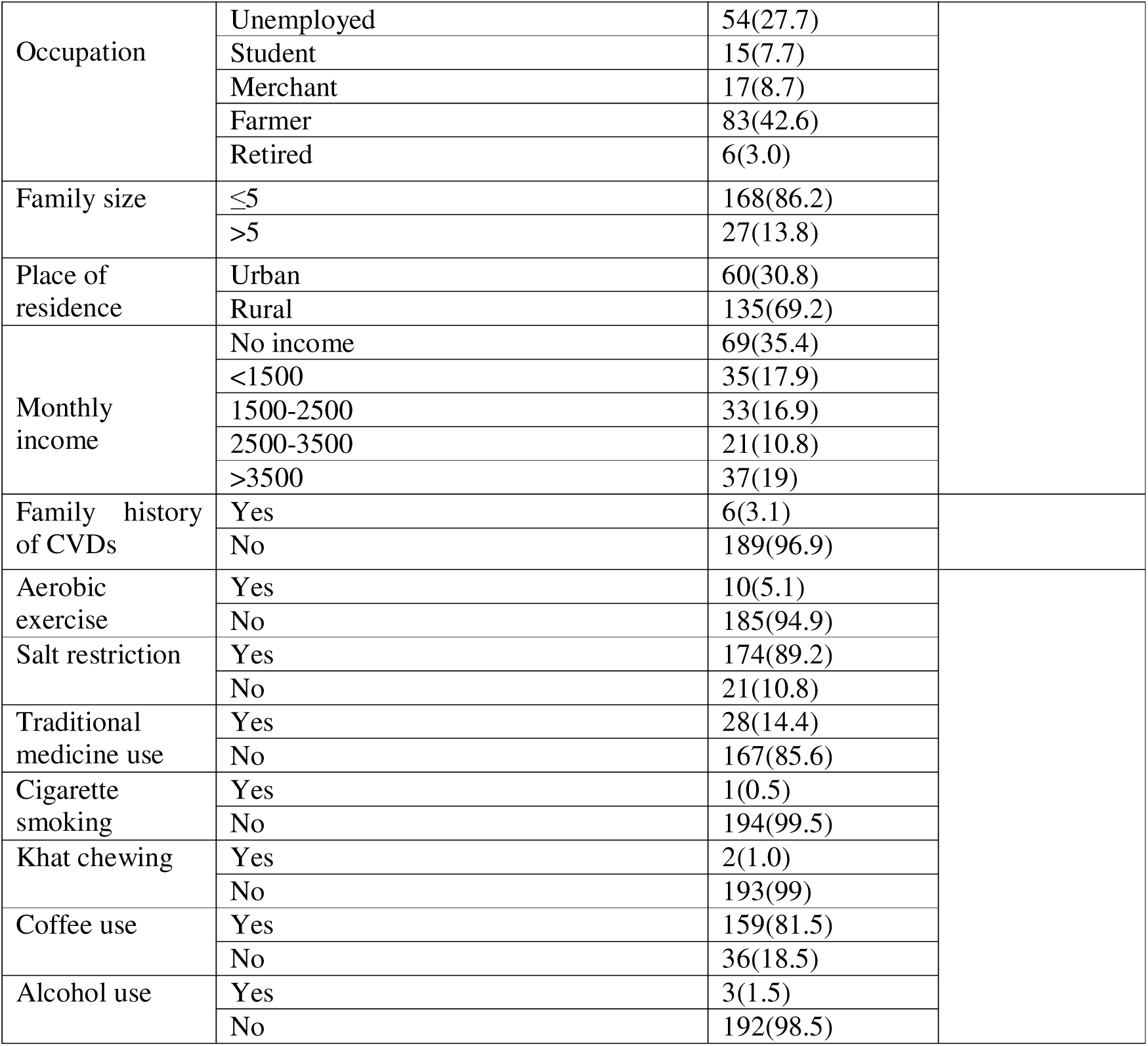
Sociodemographic characteristics of adult ambulatory patients with CVDs attending the chronic care unit at WSUCSH, from May to July, 2022 (n=195)

### 3.2 Clinical characteristics

#### Disease related variables of study patients

The mean duration of cardiovascular disease since diagnosis was 2.26 ± 0.752 years. From the study participants about 61(31.3%) had history of hospitalization while on treatment and majority 39(20%) had only one hospitalization. The most frequent comorbid conditions among the study subjects were infectious type 35 (32.8%) followed by dyspepsia secondary to PUD 15 (14%). A total of 107 comorbidities were identified from 79 study subjects in which, only one comorbidity type was identified from 60(76%) patients, two comorbidity types in 16(20.2%) patients and greater than two comorbid types was in 3(3.8%) of patients *(Figure 2).* The most common cardiovascular diseases encountered during the data collection period were hypertension 116(59.5%) and congestive heart failure 92(47.2%). The average BP levels of patients calculated from at least two consecutive values showed that 84(43.2%) of patients had poor BP control. *(Table 2)*

**Figure 2.**
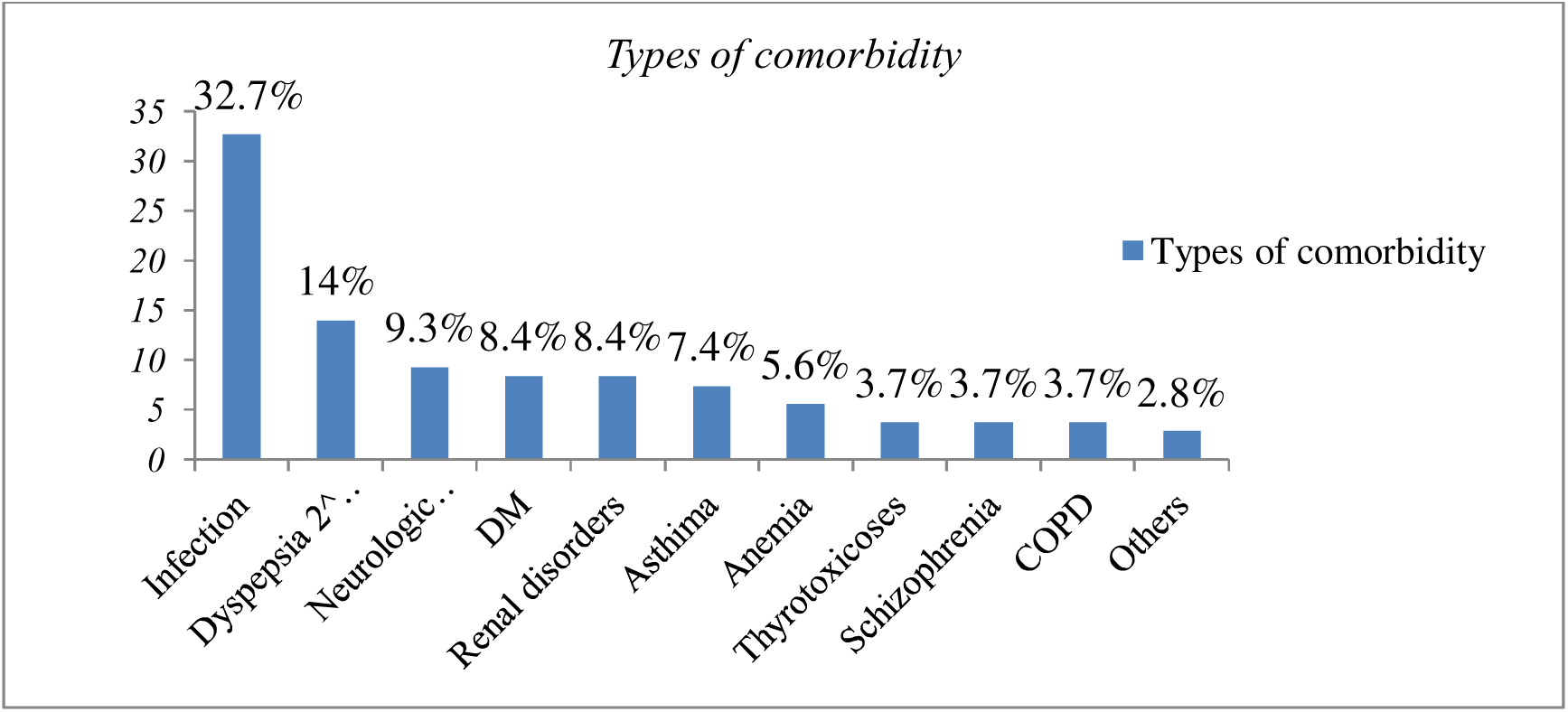
Type of comorbidity of study subjects at WSUCSH, Southern, Ethiopia, from May to July, 2022, (N=107). N.B:-Others: CLD and Nephrotic nephritic syndromes.

**Table 2:** Disease related variables among CVD patients in WSUCSH, Southern Ethiopia, from May 9 to July 8, 2022, (N=195)

| Disease related variables | Category | Number (%) | Mean ± SD |
| --- | --- | --- | --- |
| Duration of CVDs | <1 year | 36(18.5) | 2.26 ± 0.752 |
|  | 1 years | 72(36.9) |  |
|  | >1 year | 87(44.6) |  |
| Duration of therapy | <1 year | 35(17.9) |  |
|  | 1years | 71(36.5) |  |
|  | >1years | 89(45.6) |  |
| Previous Hospitalization | Yes | 61(31.3) |  |
|  | No | 134(68.7) |  |
| No of hospitalization | One times | 39(20) | 1.44 ± 0.696 |
|  | Greater than one | 22(11.3) |  |
| Reasons for hospitalization | Drug discontinuation | 7(3.6) |  |
|  | Comorbidity | 18(9.2) |  |
|  | Disease exacerbation | 21(10.8) |  |
|  | Drug discontinue& disease exacerbation | 15(7.7) |  |
| Presence of comorbidity | Yes | 79(40.5) |  |
|  | No | 116(59.5) |  |
| Types of CVDs | HTN | 116(59.5) |  |
|  | CHF | 92(47.2) |  |
|  | ACS | 4(2.0) |  |
|  | IHD | 10(5.2) |  |
|  | Stroke | 14(7.2) |  |
|  | VTE | 4(2) |  |
|  | AF | 23(11.8) |  |
| Ethology of CVDs | CRVHD | 33(16.9) |  |
|  | HHD | 35(17.9) |  |
|  | IHD | 13(6.7) |  |
|  | DCMP | 15(7.7) |  |
|  | Others | 14(7.2) |  |
| Average BP – measures | 90/60 – 130/80mmHg | 105(53.8) |  |
|  | Above 130/80 | 84(43.2) |  |
| Average glycaemic value | Less than 70 | 6(3.1) | (hypoglycaemia) |
|  | 70 – 130(good) | 50(25.6) |  |
|  | Above 130(poor) | 5(2.5) |  |
| Lipid profile | Normal | 15(7.6) |  |
|  | Dyslipidaemia | 20(10.2) |  |
|  | Not available | 160(82.2) |  |
| RFT | Normal | 121(62) |  |
|  | Impaired | 38(19.5) |  |
|  | Not available | 36(18.5) |  |
| LVEF | Not available | 43(22) |  |
|  | EF( <35) | 96(49.2) |  |
|  | EF (40—55) | 45(23) |  |
|  | EF (55—70) | 32(16.5) |  |

#### Medication related variables of study patients

About 91(46.7%) of the patients had polypharmacy. The mean number of drugs per day was 2.65±0.5 per patient. Most of the patients reported that they didn‘t experience drug allergy, only three patients (1.5%) experienced allergy, of which doxycycline accounts of (0.5%) and ceftriaxone accounts of (1*%) (Table 3)*.

**Table 3:** Medication related variables among CVD patients in WSUCSH, Southern Ethiopia, from May to July, 2022, (N=195)

| Medication related variables | Category | Frequency (%) | Mean + SD |
| --- | --- | --- | --- |
| Source of medication | Payment | 67(34.4) |  |
|  | Free | 94(48.2) |  |
|  | Both | 34(17.4) |  |
| Poly pharmacy | No | 104(53.3) | 2.65±0.5 |
|  | Yes | 91(46.7) |  |
| Drug allergy | Yes | 3(1.5) |  |
|  | No | 192(98.5) |  |
| Drug responsible for allergy | Doxycycline | 1(0.5) |  |
|  | Ceftriaxone | 2(1) |  |
| MMAS-8 | ≥8(good) | 154(79) |  |
|  | 6 – 8(medium) | 16(8.2) |  |
|  | <6(poor) | 25(12.8) |  |
| Narenjos ADR scale | ≥9(definite) | 1(0.5) |  |
|  | 5-8(probable) | 5(2.6) |  |
|  | ≤0(doubtful) | 189(96.9) |  |

**Table 4:** Types of the identified DTPs among CVD patients in WSUCSH, Southern Ethiopia, from May 9 to July 8, 2022, (N=168)

| Categories of DTPs | No of DTPs (%) |
| --- | --- |
| Unnecessary drug therapy | 20(11.9) |
| Need additional drug therapy (Unmet need) | 69(41) |
| Ineffective drug therapy | 15(8.9) |
| Dose too low | 0(0) |
| Adverse drug effects | 19(11.4) |
| Dose too high | 4(2.4) |
| Non – adherence | 41(24.4) |

A total of 640 medications were used. Commonly prescribed drug classes were diuretics (21.4%), CCB (15.9%), spironolactone (15.6%)*(Figure 3)*.

**Figure 3:**
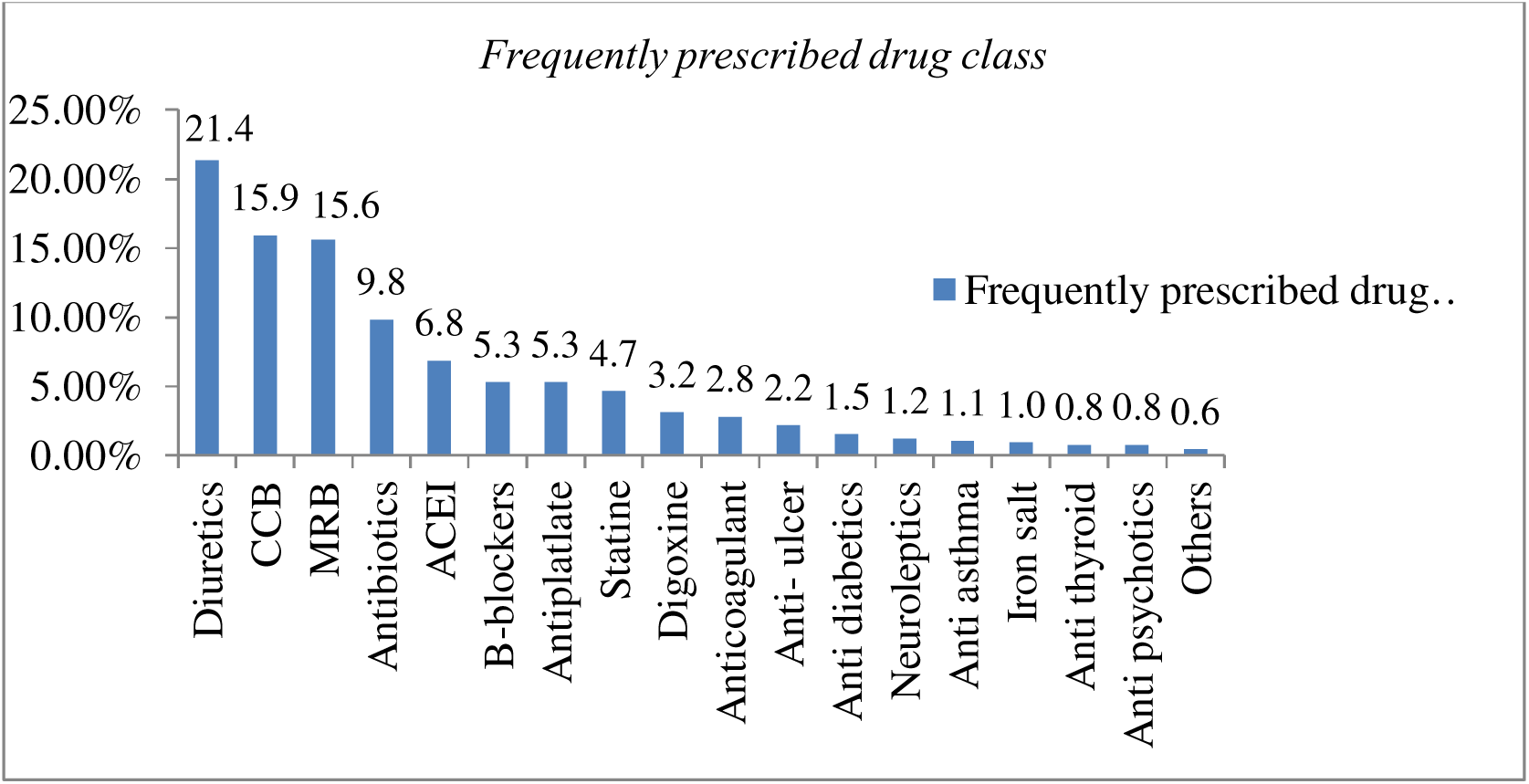
Frequently prescribed drug classes in CVD patients attending at chronic care unit of WSUCSH, Southern Ethiopia, from May to July, 2022 N= (640) **N.B:** Others: (anti-TB, HAART & NSAIDs,) ACIEs: angiotensin converting enzyme inhibitors, ARBs: angiotensin receptor blockers, CCBs: calcium channel blockers, diuretics: (loop & thiazide diuretics)

### 3.3. Prevalence and predictors of Drug Therapy problems

A total of 136 patients had one or more DTPs. The prevalence of DTPs in the current study is 69.8%. There was a total of 168 DTPs identified. The mean number of DTP was 1.3±0.461 per patient. The maximum number of DTPs was three. Most of the patients (108, 79.5%) had only one DTP. Most of the indication problem was need additional drug therapy (69, 41%).

Of the seven categories of DTPs identified, unmet therapeutic need or need additional drug therapy was found to be the most frequent 69(41% of all DTPs) type *[Table 5].* It was found that the medical condition that requires treatment 38(22.6%) was the most common reason for occurrence of this type of DTPs. According to Validated Morisky Medication Adherence Scale (MMAS), 41(24.4%) number of patients were not adhering to their medication giving non-compliance as the second most frequent type of DTPs. Majority 14(8.4%) of non-adherent patients reported that forgetting to take their medication followed by Drug unavailability & affordability 11(6.6%) were the common reasons for their non-adherence *[Table 6].* Unnecessary drug therapy 20(11.9%) was found to be the third frequent DTP category.

**Table 6:**
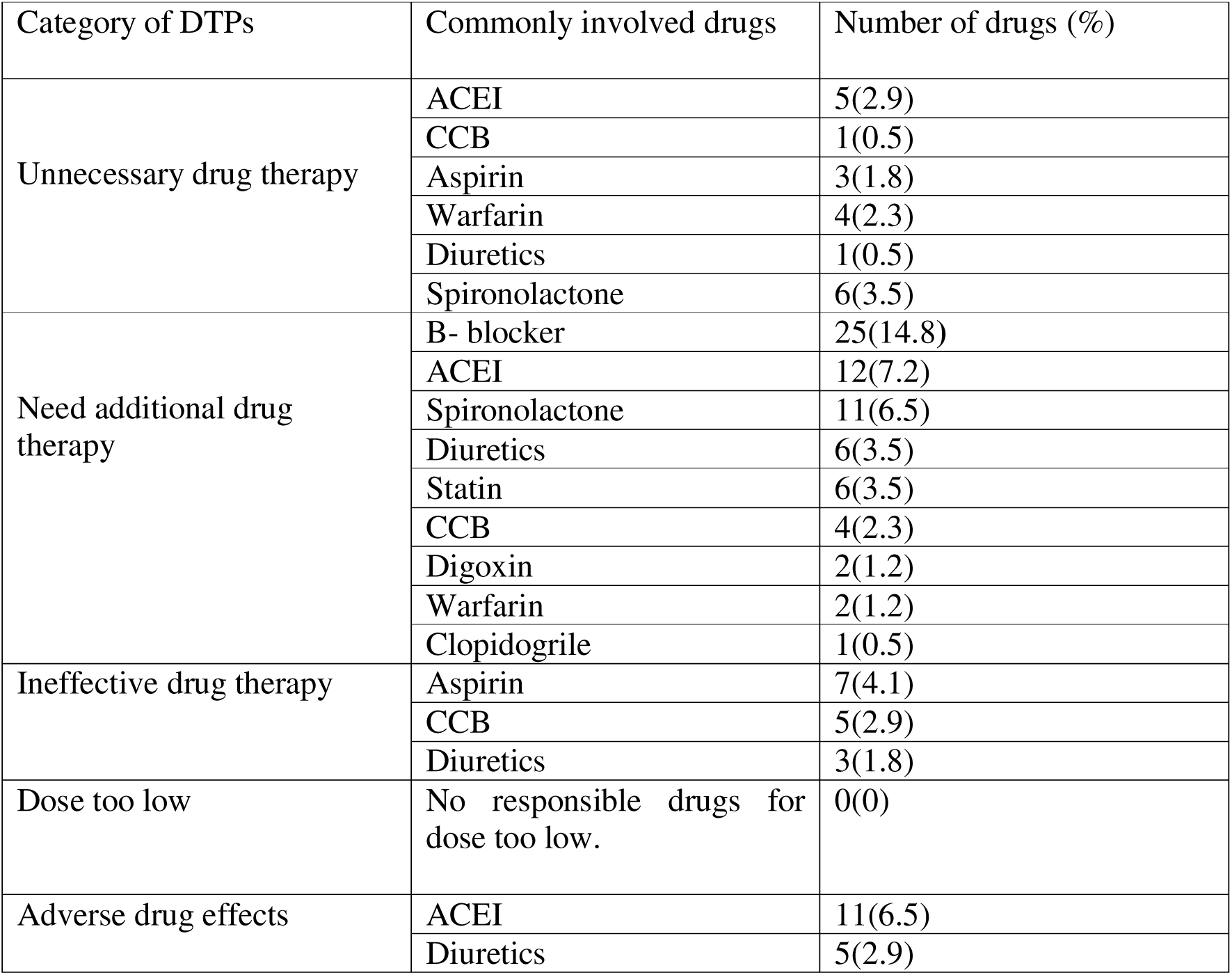
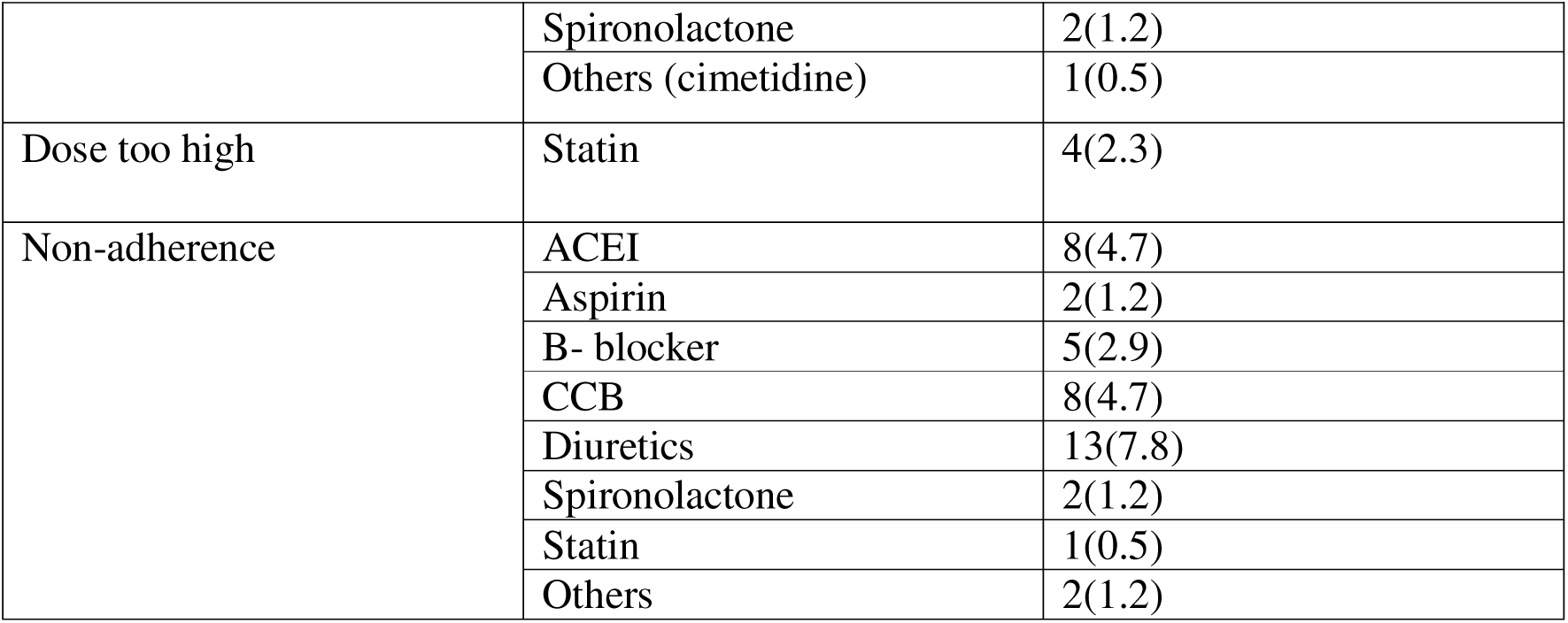
Drugs &classes of drugs involved in each category of DTPs among CVD patients at WSUCSH, Southern Ethiopia, from May 9 to July 8, 2022 (N=168)

The most frequent drug class involved in DTPs was beta blockers followed by Diuretics, ACEI, MRA, and CCBs, antiplatelet, Statin, anticoagulants and cardiac glycosides. The common indication for need additional drug therapy was cardiovascular disease with compiling indication (for example hypertension with heart failure*) [Table 7]*.

**Table 7:**
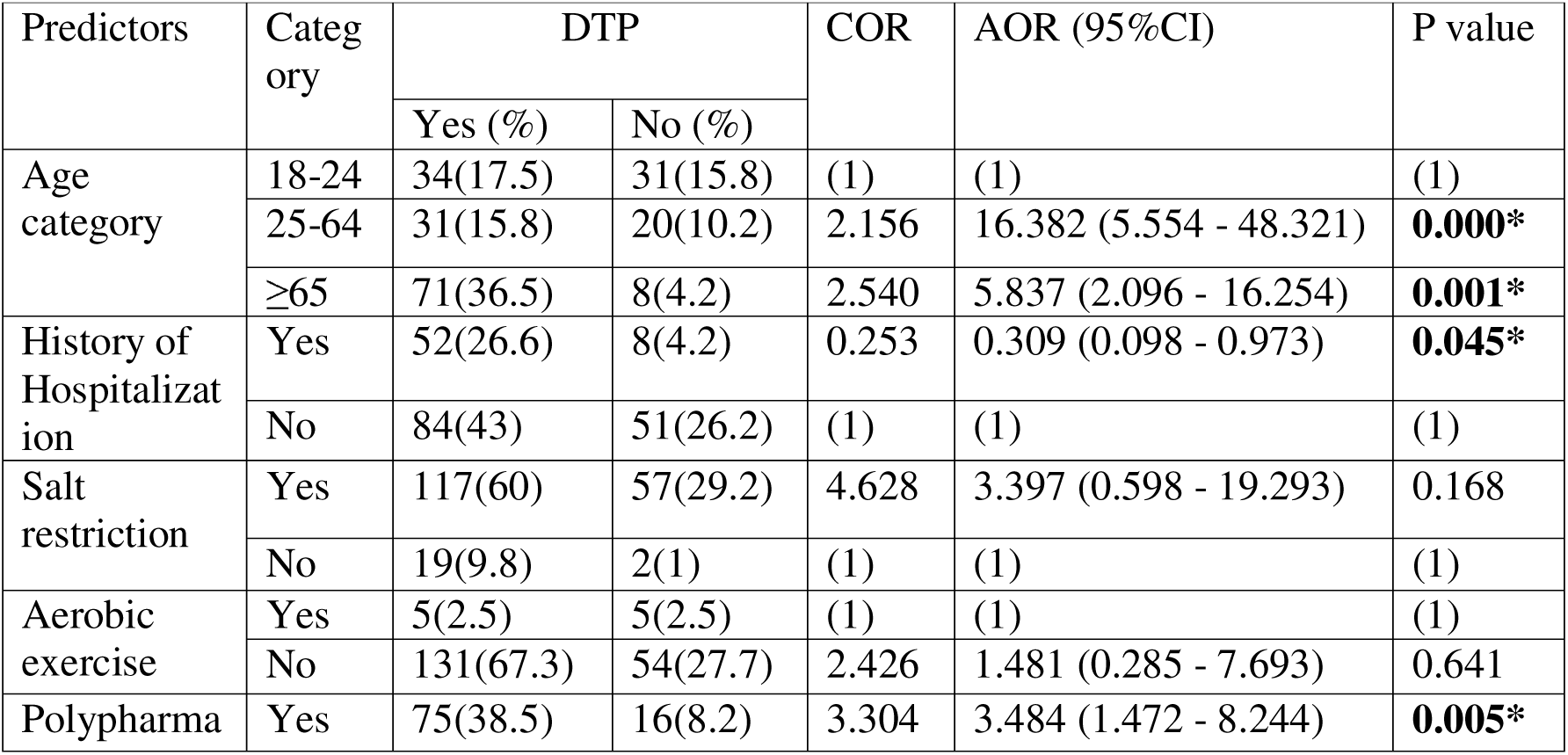
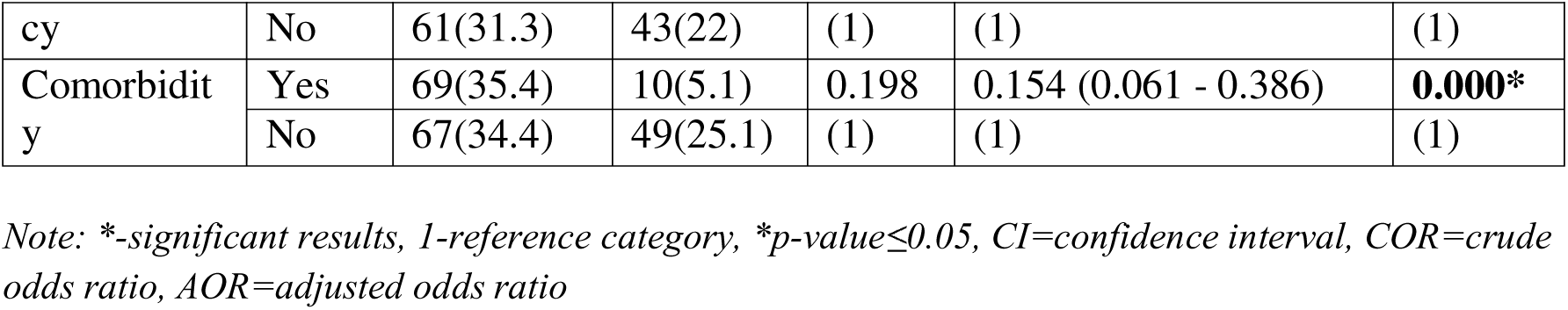
Results of multiple logistic regressions for predictors of DTPs among the study subjects in WSUCSH, southern, Ethiopia, from May 9 to July 8, 2022 (N=195)

Multivariable logistic regression analysis showed that the likely hood of having DTPs increases as age of respondents increases. Patients with in the age range of 25-64 years were 16.382 times more likely to have DTPs [AOR=16.382, 95%CI=(5.554 –48.321)] whereas those above 65 years old were 5.837 times more likely to have DTPs compared to those less than 25 years old[AOR=5.837,95%CI=(2.096 –16.254)] (P value = 0.000).Similarly, it was also found that patients who were taking more than five medications per day were about 3.484 times more likely to have DTPs [AOR=3.484, 95%CI=(1.472 – 8.244)] compared to those who were taking less than or equal to five medications (p-value of 0.005). Patients with comorbidities showed significantly reduced odds of DTPs (AOR=0.15, 95% CI: 0.06–0.39), while prior hospitalization was independently associated with lower DTP likelihood (AOR=0.31, 95% CI: 0.10–0.97). *[Table 8]*.

## 4. Discussion

This study identified that, most of the patients 136 (69.7%) with a total of 168 numbers of DTPs were identified, which is almost similar to the study done in Gebretsadik Shawo General Hospital, Bonga, Ethiopia(21). The study in FHRH showed that there were 105 DTPs(22). On the other hand a study in Jimma University Specialized Hospital (JUSH) internal medicine ward showed, 149 DTPs(23). The variation in our study can be due to inclusion of more numbers of patients with comorbidity which could increase the possibility of number of medications, clinical knowledge of investigator(s) and differences in the study method and setting may also affect DTPs. The average number of DTP in this study was 1.23±0.6 per patient but studies in FHRH and JUSH showed 1.38±0.8(24) and 3.014 (23) DTPs per patient. This is because the numbers of patients with DTP in this study are relatively higher than the two studies.

The major DTP type was unmet condition which is similar to a study in Los Angeles (25) and it was also common problem in Ethiopia FHRH (90.69%) (24), in JUSH internal medicine ward (83%)(23) and Adama(76%). While in China efficacy and safety DTP types were the common(26).This difference might be ascribed to factors such as the level of knowledge of health professionals among these health facilities regarding the appropriate indication of drugs, the proper selection of drugs as per cases observed, and issues related to comorbidities as well as the patient’s condition. Relatively lower number of safety related problems (13.7%) were found in this study than in JUSH (45.83%) (23) and by Mekonnen AB (31.39%)(27). This can be due to less types of safety related issues covered with this study including dosage too high and contraindications. Unlike Belo Horizonte(28) and European studies(29), the highest percentages of safety related issues encountered were contraindication(12.88%) and inappropriate frequency(4.90%).In Belo Horizonte the common safety issue was adverse drug events while in European studies it was dosage too high(27.5% and 13.75%) respectively. This primarily depends on patient and drug related factors.

Around 41(24.4%) number of patients were non-compliant. Forget taking the medication was the major causes for non-compliance issue; this can be due to patient factors. Even though different measurement of adherence was used, 27% of patients in Nigeria and 29% of patients in JUSH research were non-compliant. Unlike this, studies in USA (8%), Brazil (11%) and Venezuela (13%) showed relatively lower rates of non-compliance. This could be due to difference in health care service of the hospitals and majority of study patients were having educational level of non-formal education, below secondary school and age difference might also affect adherence of patients to their medications.

The most common drugs/groups of drugs responsible for the existence of DTP in this study were B-blocker, diuretics and ACEIs. This finding agrees with the study done in Jimma University Specialized Hospital, a hospital-based prospective observational study among heart failure patients, a hospital based interventional study among heart failure patients in Barcelona and similarly study done in Taiwanese heart failure patients.

With regard to variables influencing the occurrence of DTPs among patients with CVD; multiple regression analysis indicated that age, polypharmacy, history of hospitalization and comorbidity independently predicted the occurrence of DTPs in this study. The use of multiple drugs was significantly associated in both binary logistic and multivariate logistic regressions. in multivariate logistic regression analysis, those patients who received greater than five drugs were 3.484 times more likely to experience DTPs than those who were using five or fewer drugs. This finding is in agreement with the study done in Jordan where the number of medications used were strongly linked to treatment-related problems(30). As well as the study conducted in the northwest of Ethiopia in which the number of medications per patient is associated with the risk of DTPs (31). In addition, the finding of our study is also in agreement with another study conducted in the southwest of Ethiopia in which the use of more than five drugs per day per patient was found to be a predictor of the occurrence of DTPs.

It was also found that the likely hood of occurrence of DTPs was increasing as age of the respondents’ increases. Patients found within age range of 25-64 years were about sixteen times more likely to develop DTPs than those below 24 years whereas the factor increased to five times for elderly patients (age above 65 years) Association of advanced age with DTPs has prone scientific ground as it results in multiple disease conditions requiring multiple medications. This finding is consistent with a study in Jordan(32) and in Taiwan(33). But a study done in Ethiopia, Hiwot Fana Specialized University Hospital(34) showed no significant association of age with DTPs. The discordance could be due to difference in study patients, and due to most of the study participants in this study were elderly patients.

Notably, we identified a protective association between comorbidity as well as prior hospitalization and DTP occurrence. While this finding contrasts with some literature, it may reflect the benefits of structured medication reconciliation during hospital discharge and enhanced follow-up care. Potential explanatory factors include: – comprehensive medication review during hospitalization, improved patient education during inpatient stays and more rigorous post-discharge monitoring protocols.

## 5. Conclusion

In conclusion, this study highlights significant medication management challenges among ambulatory CVD patients, with a majority experiencing drug therapy problems. The most prevalent issues were untreated conditions and medication non-adherence, particularly affecting younger patients and those on multiple medications. While some clinical factors appeared protective, the high prevalence of therapy gaps and cardiovascular risk underscores the urgent need for enhanced medication review processes and coordinated care strategies to optimize treatment outcomes in this vulnerable population.

## Limitations of the study

This study has some limitations. Its cross-sectional design assessed only current DTPs without follow-up or interventions. Non-adherence may be overestimated due to self-reporting bias in the Morisky scale. Missing data (e.g., HbA1c, BMI, RFT, LFT, lipid profiles) limited analysis. Additionally, being a single-centre study, broader multi-centre research is needed for generalizability.

## Data Availability

All data produced in the present study are available upon reasonable request to the authors

## Acknowledgement

We would like to acknowledge the nurses, pharmacists, and physicians who were involved in data collection and expert decision in DTP identification. Our gratefulness extended to the CVD patients for their eager involvement in the study.

